# Mapping the effectiveness of the community tuberculosis care programme: A systematic review

**DOI:** 10.1101/2022.08.22.22279091

**Authors:** Gabalape Arnold Sejie, Ozayr H Mahomed

**Affiliations:** Discipline of Public Health Medicine, University of KwaZulu, Natal, Durban, South Africa

## Abstract

**Background:** Tuberculosis (TB) is a major public health problem throughout the world particularly in resource limited countries. In light of the global urgency to improve TB care, the World Health Organisation emphasize the importance of taking into consideration the journey of a TB patient through a series of interlinked settings and facilities. One of these is decentralising TB care beyond health facilities and harness the contribution of communities through provision of effective community-based directly observed therapy (DOT) to TB patients at greatest socio-economic risk. A systematic review was conducted to map previously conducted studies to identify existing community TB implementation models, their effectiveness on cost and treatment outcomes.

**Methods:** Systematic search through various electronic databases electronic databases; Medline/PubMed, EBSCO (PsycINFO and CINAHL) and Cochrane libraries was performed between the year 2000 and 2021. We used the following free text search terms Tuberculosis, Community tuberculosis, cost effectiveness and treatment outcomes for this purpose. Their quality was scored by ROBINS-I and ROB 2.

**Results:** A total of 6982 articles were identified with 36 meeting the eligibility criteria for analysis. Two observational studies in low-and middle-income countries reported comparable video observed treatment completion rates to in-person directly observed therapy (0.99-1.47(95% CI 0.93-2,25) with one randomised control trial in a high-income country reporting an increased video observed treatment success rate to standard care (OR 2.52, 95% CI 1.17-5.47). An incremental cost saving ranged was $1391-$2226. Electronic medication monitors increased the probability of treatment success rate (RR 1.0-4.33 and the 95% CI 0.98-95.4) in four cohort studies in low-and middle-income countries with incremental cost effectiveness of $434. Four cohort studies evaluating community health worker direct observation therapy in low-and middle-income countries showed treatment success risk ratio ranging between 0.29-3.09 with 95% CI 0.06-7.88. (32,41,43,48) with incremental cost effectiveness up to USS$410 while four randomised control trials in low-and middle-income countries reported family directly observed treatment success odds ratios ranging 1.03-1.10 95% CI 0.41-1.72. Moreover, four comparative studies in low-and middle-income countries showed family directly observed treatment success risk ratio ranging 0.94-9.07, 95% CI 0.92-89.9. Lastly four Short Message Service trials revealed a treatment success risk ratio ranging 1.0–1.45, 95% CI fell within these values) with cost effectiveness of up to 350I$ compared to standard of care.

**Conclusions:** This review illustrates that community-based TB interventions such as video observed therapy, electronic medication monitors, community health worker direct observation therapy, family directly observed treatment and short Message Service can substantially bolster efficiency and convenience for patients and providers thus saving costs and improving clinical outcomes.

## Background

Tuberculosis (TB) is a notable health risk to current and future global health, with millions of people continuing to fall sick with the disease each year. In 2021, World Health Organisation (WHO) projections indicated TB as the 13^th^ leading cause of death and the second deadliest infectious killer after COVID-19.(1) Globally, an estimated 9.9 million people fell ill with TB in 2020, a number that has been relatively stable in recent years.(1) The burden of disease varies enormously among countries, with the global average being around 127 cases per 100 000 population. (1)TB affects people of both sexes in all age groups however the highest burden is in men, who accounted for 56% of all TB cases in 2020. Geographically, most TB cases in 2020 were in the WHO regions of South-East Asia (43%), Africa (25%) and the Western Pacific (18%), with smaller percentages in the Eastern Mediterranean (8.3%), the Americas (3%) and Europe (2.3%).(1) African region had the most TB burden countries with highest TB incidence of 237 cases per 100 000 population.(1) The proportion of TB cases coinfected with HIV was highest in countries in the WHO African Region, exceeding 50% in parts of southern Africa.(2)

The global TB treatment coverage down from 72% in 2019 to 59% in the year 2020. Among the six WHO regions, treatment coverage was highest in Europe with a best estimate of 69% and lowest in the Eastern Mediterranean with a best estimate of 52%. (2) The treatment success rate for the new and relapse cases treated in the 2017 cohort globally was 85%. (1) Among the six WHO regions, the highest treatment success rates in 2017 of 91%, were in the WHO Eastern Mediterranean Region and the Western Pacific Region. The lowest rates were 76% in the WHO Region of the Americas and 78% in the European Region. (1) There were an estimated 1.3million TB deaths among HIV-negative people in 2020, and an additional 214 000 deaths among HIV positive people with about 83% of TB deaths among HIV-negative people occurring in the WHO African Region and the WHO South-East Asia Region.(1) The global reduction in the total number of TB deaths between 2015 and 2020 was 9.2%, about one quarter of the way to the End TB Strategy milestone of a 35% reduction by 2020.(2)

The WHO End TB Strategy envisions a TB-free world by the year 2035(3). In light of limitations inherent in prevailing tuberculosis care and the global urgency to improve TB care, the WHO emphasize the importance of taking into consideration the journey of a TB patient through a series of interlinked settings and facilities(4).One of the ways to do this is by decentralising TB care beyond health facilities and harness the contribution of communities through provision of effective community-based directly observed therapy (DOT) to TB patients at greatest socio-economic risk.(3) These patient centred community-based care models are considered necessary to increase the capacity of the public health system to provide treatment to more TB patients and may address patients’ needs more successfully as care closer to home is easier to access, convenient, allows family support and eliminates long and costly trips to a centralized primary health facility(5)(6). However, despite the clear need, the documented cost-effectiveness of community-based TB activities and the tremendous efforts that have been expended in recent years, the program is not yielding expected results. Treatment success rates in many countries are declining and remaining below global targets, resulting in increased health care expenditure and poor quality of life to the victims (1). Projected trends indicate that a substantial strengthening of efforts to reduce TB incidence is needed if the WHO End TB strategy is to be met in countries of high TB incidence(7). Whereas frameworks to accelerate the reduction in TB incidence can be reached by expert consensus, it is fundamental that these are underpinned by a robust and up-to-date evidence base for the effectiveness of specific interventions. Such an evidence base also allows for harmonization of best-practice approaches to TB control (7). The purpose of the systematic review is to aid the research in identifying previously conducted studies to identify what models of community TB implementation exists, their cost effectiveness and effectiveness on patient outcomes. The systematic review will also assess the disadvantages and advantages of the different models of implementation in relation to cost effectiveness in low-and medium-income countries (LMIC). This review findings aspire to contribute to the knowledge base in the area of community TB care implementation, inform policy, planning and guide future research.

## Methods

The study was analysed and reported in accordance with the Cochrane systematic review guidelines and the Preferred Reporting Items for Systematic reviews and Meta-Analyses (PRISMA) to include constituents that resonate with the underpinnings of the systematic review methodology(8)(9). Investigators developed the systematic review protocol including the eligibility criteria and the Data abstraction tool. No formal protocol was published for this systematic review

### Literature search strategy

A comprehensive search strategy was developed using appropriate key words, Medical Subject Headings (MeSH) and free text terms to maximize the retrieval of potentially relevant studies. The search was conducted across various electronic databases between the year 2000 and 2021: Medline/PubMed, EBSCO (PsycINFO and CINAHL), Cochrane libraries, EMBASE and WHO Regional Databases; Keywords used to search for these databases included but not confined to: Tuberculosis, Community tuberculosis care, Implementation models, cost-effectiveness and treatment outcomes. During the search, keywords were separated by Boolean terms (AND, OR, NOT). We complemented these database search results with ‘grey literature,’ including hand-searched bibliographies to identify any studies which may have been missed by the above search strategies. Studies obtained through database searches were exported to Endnote library for further abstract and full article screening respectively. The Endnote library “find full text” option was used to automatically download PDFs of exported studies. A global search approach was utilized then contextual interpretation for low middle income countries.

### Screening and eligibility determination

Using a two-reviewer system (with consensus for disagreements and conferral with a 3rd party adjudicator if a consensus was unable to be reached), all articles identified through the above literature search were screened by reviewing the title and abstract to remove all articles that clearly did not meet the eligibility criteria. The full text of the remaining articles were reviewed by two reviewers, any ensuing discrepancies were resolved by discussion or the involvement of the third reviewer if a consensus was not reached by the first two reviewers. In accordance with recommendations by Levac et al, (10) after reviewing every batch of 20 to 30 publications, the reviewers meet to resolve any conflicts and ensure consistency with the research question and purpose. To capture and present the screening process, the Preferred Reporting Items for Systematic and Meta-Analyses flow diagram in **Fig 1** was used(9)(11).

**Figure 1:**
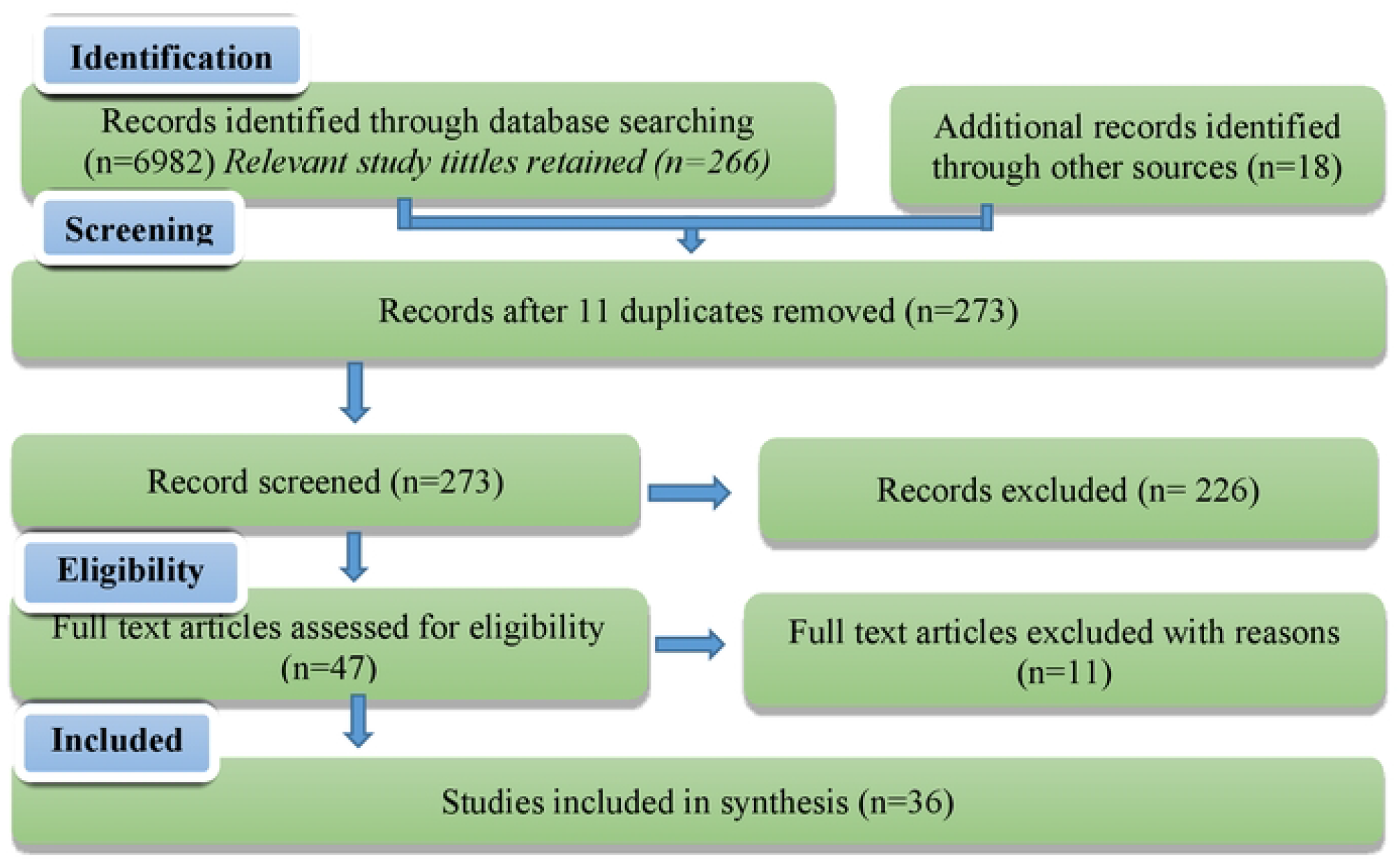
Preferred Reporting Ite1ns for Systematic and Meta Analyses flow diagra1n. (Adapted fro1n Moher et al).(11)

Eligibility was based on the following inclusion and exclusion criteria:

### Inclusion criteria

- Evidence published in English.
- Publications between 2000 and 2021
- Literature with substantial focus on community TB implementation models including peer-reviewed journal articles, systematic reviews, scoping reviews, meta-analysis and rapid reviews, government and non-governmental organization reports and academic dissertations.
- Research focusing on community TB implementation models in low-income and middle-income countries and whose conclusions and discussion demonstrate transferable findings to Botswana settings.
- All study designs were considered including qualitative, quantitative and mixed-methods studies.

### Exclusion criteria

- Studies were excluded if the intervention was purely facility-based
- Articles that are not in English were excluded as the researcher will not assess them due to the language barrier

### Methodological quality assessment

We assessed the risk of bias in randomised control trials (RCTs) in this review using the Cochrane risk-of-bias tool for randomized trials (RoB 2) according to the following domains: bias arising from the randomization process; bias due to deviations from intended interventions; bias due to missing outcome data; bias in the measurement of the outcome; and bias in the selection of the reported result. (12) Cluster randomised trials were assessed using the risk of bias for cluster randomised studies(8).

Observational studies were assessed using the Risk Of Bias In Non-randomised Studies-of Interventions (ROBINS-I) tool. The ROBINS-I assesses four broad areas: confounding, selection bias, information bias, and reporting biases. (13) The overall quality of the evidence for the primary outcome was assessed with the adapted GRADE approach(8). Domains that may decrease the quality of the evidence are study design and implementation (risk of bias), inconsistency (heterogeneity), indirectness (inability to generalize), imprecision (insufficient or imprecise data), and publication bias across all studies that measure that particular outcome. The quality of the evidence on a specific outcome is based on the performance against six factors: study design, risk of bias, consistency, and directness of results, the precision of the data, and publication bias across all studies that measured that particular outcome. Two reviewers appraised each study independently and disagreements were resolved through discussion with a third reviewer.

### Charting the data and collating, summarising, and reporting the results

A comprehensive data abstraction format in Microsoft Excel was developed collectively by the reviewers to extract predetermined variables. Prior to full extraction, the tool was tested with four articles and refined. Structuring this Excel sheet database involved selecting and defining data categories and subcategories. It was secured online so that involved reviewers will have access and can make updates freely. Bibliographic details, study design, intervention(s), comparison(s), measures of effect (risk ratios, or odds ratios with respective confidence interval) outcomes, study setting and conclusions for the primary and secondary outcomes of interest were extracted. The geographic origin of the papers was categorized according to the World Bank country classification by economic level which includes low-and middle-income countries and high income(developed) countries. This dataset was populated from each selected paper. This step was done iteratively as more familiarity of literature is gained and revisions done as appropriate. This was purposively done to keep track of the studies included and excluded during the charting process of the systematic review. Two independent reviewers did data charting. The extracted data were extrapolated into a data charting form in a Microsoft Excel file depicting: existing community TB implementation model as it relates to study designs used, type of models, their effectiveness on treatment outcomes and cost effectiveness. The data was analyzed using quantitative approach to address the main aim and the specific study questions. Further to this, the study team scrutinized the meanings of the findings as they relate to overall purpose of the study, discuss the implications for future research, practice and policy. The goal of the systematic review is to provide an overview of the available literature, so all studies were included regardless of quality assessment outcome. Information, including predetermined variables are summarised descriptively. See **Table 1** for the data extraction form.

## Results

### Results of the search

A total of 6982 references was identified from the bibliographic search. Of this, screening of titles yielded 284 studies. After removing duplicates, we identified 273 potentially relevant references; 226 were excluded based on title and abstracts, leaving 47 studies that were acquired in full text or study report with available information for further evaluation. After conducting a full-text review, thirty-six studies were included in our systematic review. A hand-search of references of the included studies revealed no further relevant publications. We used PRISMA checklist for assessment of meta-analysis guideline compliance. **Fig 1**

### Study characteristics

The thirty-six included studies were published between 2000 and 2021. They consisted of eleven RCTs(14,15,24,16–23), four cluster randomised control trials(25–28), nineteen seventeen cohort studies(29,30,39–47,31–38), one records reviews(48) and one quasi trial(49). All thirty-six had a control group and provided estimates of effect: Eleven evaluated community health care worker direct observation therapy (CHWDOT)(24,27,31,38)(14,19,25,26,29,33,39), nine evaluated family direct observation therapy (FDOT) (21–23,28,37,40,41,48–50), five evaluated short Message Service reminders (SMS) (15–18,51,52), six examined video observed therapy(VOT) (20,32,34,35,43,44) and five examined the electronic medication monitors (EMM) (30,42,45–47) See **Fig 2**. Substantive descriptions of the included studies including intervention evaluated can be seen in **Table 2 and 3**

**Fig 2:**
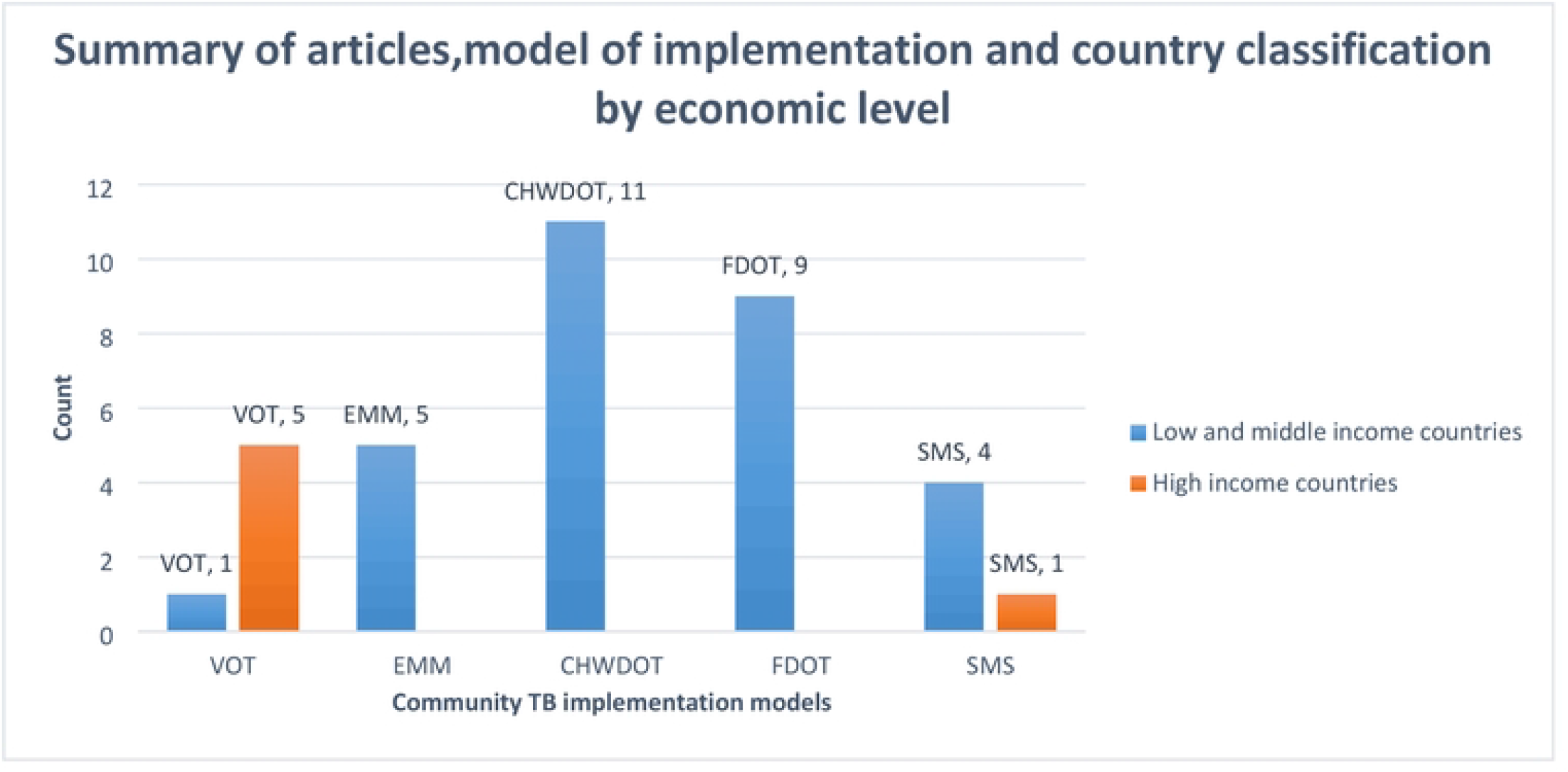
Summary of articles showing model of implementation and country classification by economic level.

Studies were excluded from the analysis for any of the following reasons: No exposure of interest, No outcome of interest, qualitative studies, articles available only in abstract form, case reports, anonymous reports, studies reporting data purely on facility-based TB, and articles that are not in English because of unfulfilled the inclusion criteria. (53,54,63,55–62)**Table 4**

### Risk of bias assessment

Quality assessment and risk of bias in the randomized trials reviewed are shown in **Fig 3**. We found low selection bias, as all eleven publications of randomized trials provided information about the processes of random sequence generation and/or allocation concealment in the studies. Overall, there was high bias of performance across the trials. Detection/outcome measurement bias was high for two trials since the assessors were aware of the intervention received by study participants. Studies varied with respect to attrition bias. We also found low reporting bias in nine of the trials. For clustered RCT a risk of bias 2 for clustered tool was used, four studies were evaluated with the overall risk of bias being low. (**Fig 4**)

**Fig 3.**
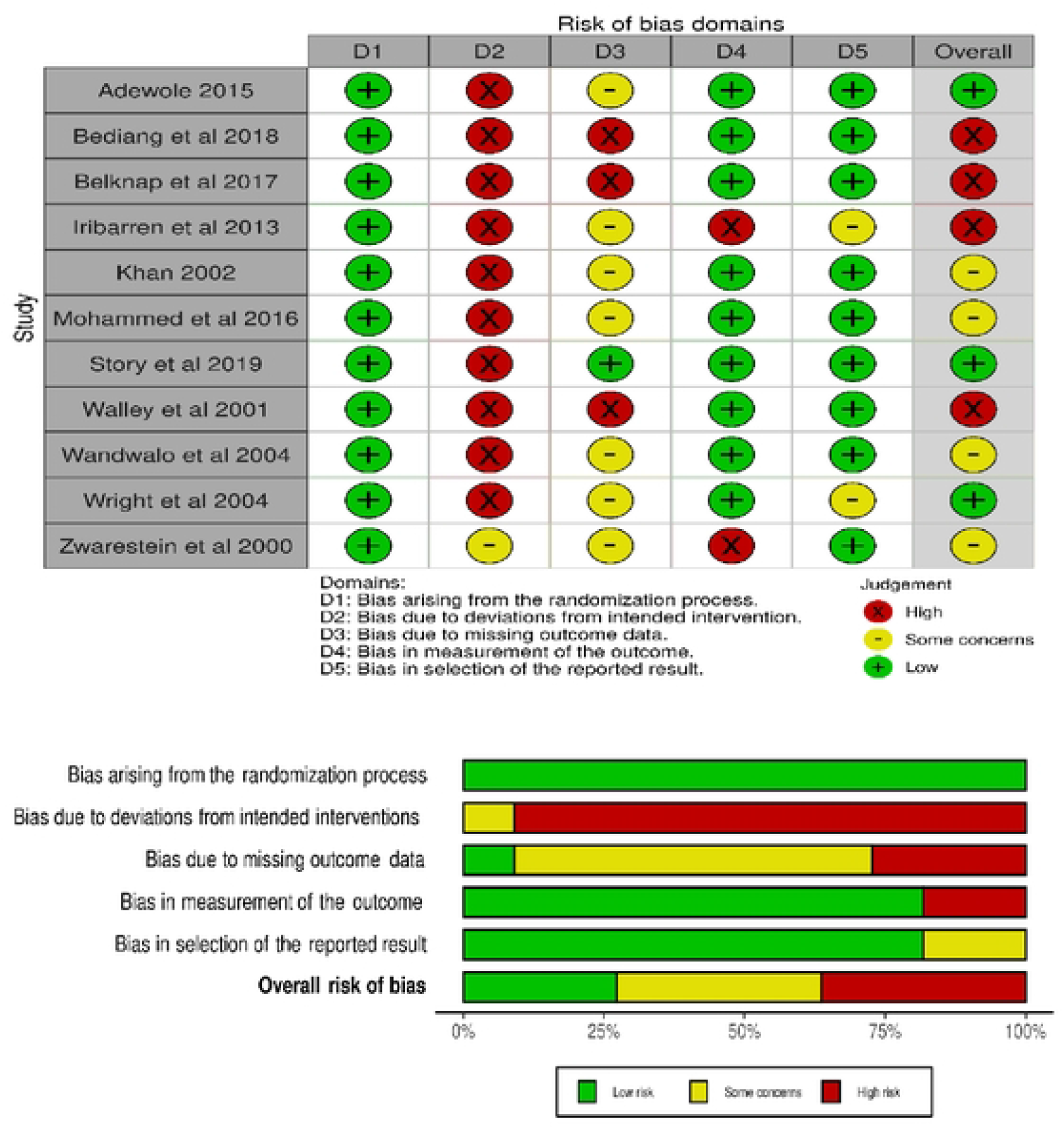
The risk of bias assessment of included papers using ROB 2 tool for randomized studies.

**Fig 4.**
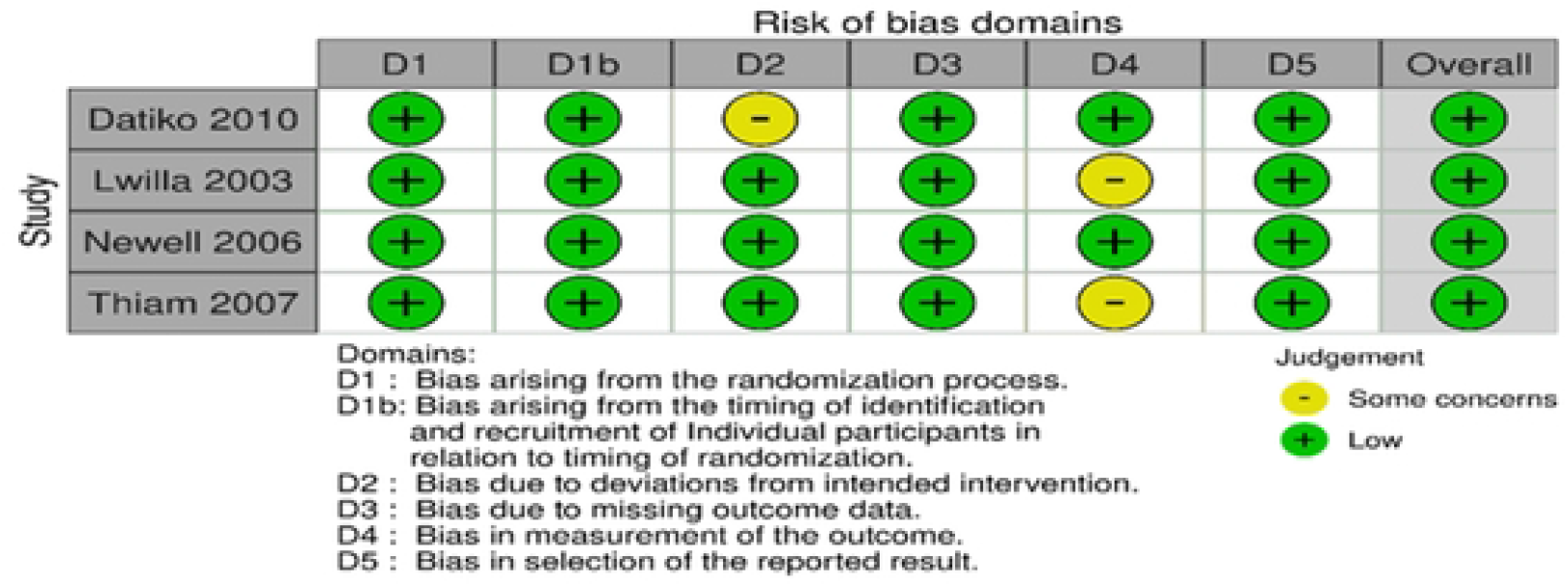
The risk of bias assessment of included papers using ROB 2 tool for cluster-randomized studies.

Twenty-one studies included were observational, methodologic quality in these studies was assessed using the Cochrane risk-of-bias in Non-randomized Studies of Interventions” (ROBINS-I) scoring system (13). ROBINS-I views each study as an attempt to simulate an ideal randomized trial, that is expected to answer a particular clinical problem. Seven domains were investigated for potential risk of introducing bias, that are judged with use of signaling questions. Overall, the risk of bias was moderate in most papers, which is understandable as most studies were non-randomized and had the retrospective design, and as such are subject to confounding and a range of other biases (**Fig 5**).

**Fig 5.**
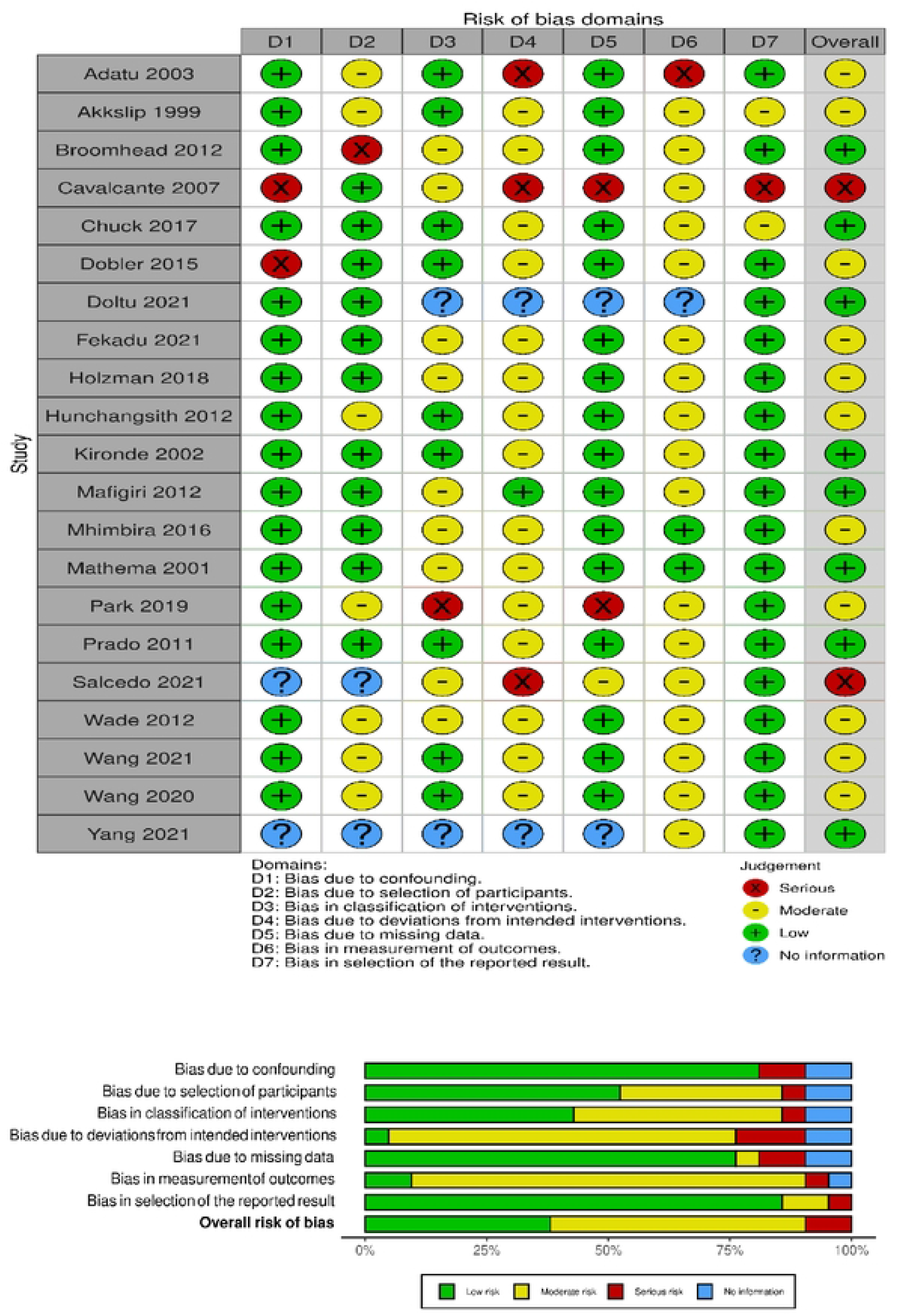
The risk of bias assessment of included papers using ROBINS-I tool for non-randomized studies.

### Existing community TB care models

In this review, we identified five models of implementation to form basis of our analysis. These models include Video observed therapy (VOT), Electronic medication monitors (EMM), Community health care worker Directly observed therapy (CHW DOT), Family-member Directly observed therapy (FDOT), and Short Message Service (SMS). Explanation of the models implementation are discussed below.

### Video observed treatment (VOT)

Six studies reported on this model, the model can be utilised both Asynchronous and Synchronous. The asynchronous video-observed therapy (VOT) application enables participants to record themselves swallowing each treatment dose and send videos for review by a DOT worker. Each recorded dose is automatically date-and time-stamped, encrypted, and uploaded to a secure server over a cellular or wireless network. Once the data is received by the server, the smartphone application deletes videos from the device to prevent unintentional disclosure of participant information and conserve device memory. Videos are stored on the smartphone in a manner that prevented viewing, editing, resending, or deleting them to protect participant privacy and ensure video fidelity. The asynchronous design allows participants to take their medications regardless of network connectivity (e.g., while traveling) because videos uploaded automatically whenever cellular or WiFi connections is established. An application status screen allows participants to see when videos were uploaded or pending(64) (34)(20). This intervention can also be accessed using a live video (Synchronous video-observed therapy), here the VOT worker and patient pre-arrange a schedule for the VOT calls. The VOT worker receives calls using a webcam equipped computer. The patient shows and names each pill in front of the camera before swallowing it. To demonstrate that the pills had been swallowed, the patients open their mouths in front of the camera and engages in conversation with the VOT worker for several minutes. If any side effects are reported, a physician will be connected by video or audio to provide medical advice. Each VOT session are documented in the electronic medical record (EMR) system. Missed VOT appointments are followed up by phone calls and, if these is unsuccessful, home visits. The VOT worker records the start and end time of each session, including the time it took to document the session, in the EMR and the VOT database.(32)(34)

### Electronic medication monitors (EMM)

Five studies reported on automated electronic devices that monitor and store adherence records combined with audible reminder alarms. This model consists of a device that attaches to the standard pill bottle or blister pack and record/sends an SMS every time the patient opens the bottle to a Web-based application indicating the patient has taken his or her medication. During each time the patients visited TB designated hospitals at the county level, doctors generate the adherence report by connecting the EMM to an offline software program in their computers. Appropriate measures will then be taken depending on number of doses missed.(45)(42)(30)

### Community health care worker Directly observed therapy (CHW DOT)

Eleven studies reported on community health care worker Directly observed (CHWDOT), the community health care worker was defined as someone living in the same village as that of the patient and observed the patient daily. In this model of implementation, the CHWDOT observers who volunteered to work are interviewed and selected by the village leaders and trained to supervise the patient’s intake of the TB medications. The CHWDOT observers are visited fortnightly by the TB health worker in charge of the nearby health facility. During house visits, the health workers monitor adherence to treatment by checking the treatment card and counting pills remaining from the patient’s monthly drug supply. During the whole course of treatment, patients under CHWDOT have to visit the health facility monthly for drug collection. Drugs are collected from a nearby health facility by a patient or her/his CHWDOT observer, depending on the condition of the patient.(24,26,27,29,31,33,38,39,65)

### Family-member Directly observed therapy (FDOT)

Nine studies reported on Family DOT strategy in which anti-TB medications are administered at home under the supervision of an adult household member. (66,67)There are no specific criteria related to education or occupation of the family member to qualify as DOT provider. If client do not want to have a family member provide DOT, then they are considered ineligible for the intervention. In such cases a Family supervisor was assigned to the client. the eligible family member who wished to become the DOT provider is given onsite training (at home) by government supervisory staff. The training focuses on the DOTS strategy, the treatment process and its duration, the role of DOT supervisors in ensuring TB treatment completion, and side effects of anti-TB medications. The family member collects drugs fortnightly from the health facility most convenient for him or her. As per national guidelines, there would be two treatment cards for each patient: the original treatment card with the community DOT provider and a duplicate treatment card at the health Centre which is updated fortnightly by the government supervisory staff. All participants are later visited by either a medical officer or a treatment supervisor assigned to their respective area. This step is taken to verify whether each client is receiving family DOT according to national operating procedures. Treatment monitoring is done by following up of the client as per national guidelines (clinical/bacteriological assessment at the end of 2 months, and at the end of treatment).(21–23,28,40,41,48,50,66,67)

### Short Message Service Reminders (SMS)

Five studies evaluated the SMS reminders Directly observed treatment, this intervention utilizes a daily medication reminder system for TB patients. Once a patient is enrolled, the system send daily SMS reminders scheduled at a time that the patient specified during enrolment. The messages include a motivational message followed by a reminder to patients to reply via SMS to indicate that they have taken their medication. Messages are sent every day throughout the full duration of patients’ treatment. To maintain the patients’ attention and interest, these messages are changed every 2 weeks. If a patient did not reply, additional reminders are sent, and the team would follow up to confirm that the messages are being received.(15–18,68,69)

### Model/modality clinical effectiveness

The measure of effectiveness was based on sputum smear results at the end of the 2nd and 6th months of treatment. Patients with at least two negative smears including the smear at the 6th month were reported as cured. Patients who finished the treatment but did not have the 6th month smear result were reported as treatment completed. We used treatment success rate (TSR)as a measure of effectiveness, which is a standard indicator used by WHO to measure programme success. TSR was calculated as the sum of the number of TB patients who were cured and the number of TB patients for which treatment was completed divided by the total number of smear-positive cases reported, expressed as a percentage.

### Video observed treatment (VOT)

Two cohort studies conducted in developed countries of Australia and United State of America in 2012 and 2017 reported a risk ratio ranging between 0.99-1.47(95% CI 0.93-2,25) for treatment completion with VOT compared with in-person DOT(32) (44). Another cohort study in the low-and middle-income country of Moldova in 2021 reported a treatment success risk ratio of 0.07 (95% CI 0.0-0.5) to VOT compared with Facility DOT(34). One RCT in a high income country of the United Kingdom conducted in 2019 reported a higher proportion of treatment completion rates with VOT compared with in-person DOT, (OR 2.52, 95% CI 1.17-5.47), but the effect on treatment completion rates was not statistically significant.(20)This results suggest that VOT is marginally effective in the low and income countries but more effective in the high income countries.

### Electronic medication monitor (EMM)

Four cohort studies from low-and middle-income countries of China, Morocco and South Africa between 2012 and 2021 evaluated electronic medication monitors against DOT and self-administered treatment (SAT). The results of these studies showed a treatment success ratio ranged from 1.0-4.33 and the 95% CI fell within 0.98-95.4 values all with no statistical significant effect on treatment success. (30,42,45,46) Suggesting that electronic medication monitor is effective on treatment outcomes especially in the countries with lower economies.

### Community health care worker Directly observed therapy (CHWDOT)

Four cohort studies conducted in low-and middle-income countries of South Africa, Uganda, Brazil, Kampala and Mongolia between the year 2002 and 2015 evaluating community health worker delivered DOT against family DOT and facility or family based DOT showed risk ratio ranging between 0.29-3.09 with 95% CI falling between 0.06 and 7.88. (31,33,38,39) Of these four studies, only one study in Uganda did not find the difference in the treatment success rate between home-based DOTS and clinic based DOTS (OR=0.29; 95% CI: 0.06 to 1.34).(39) Three RCT also from countries of the low-and middle-income(Tanzania, South Africa and Senegal) between 2000 and 2007 reported a treatment success risk ratio of 1.18-1.7(95% CI:0.1-34.5). (27)(24)(26).

### Family-member Directly observed therapy (FDOT)

Four RCT all from low and middle income countries of Tanzania, Eswatini, Nepal and Pakistan evaluated family DOT against community health care workers, self-administered treatment and facility DOT between 2001 and 2006 showed odds ratios ranging from 1.03 to 1.10 with the 95% CI falling between 0.41-1.72, all without statistically significant effect on treatment completion, success, cure rate.(21)(28)(23)(22) Four comparative studies (Quasi trial, retrospective cohort analysis and records review) still from low and middle income countries of Brazil, Thailand, Nepal and Tanzania between 2001 and 2016 showed no statistical significant effect on treatment success with risk ratio ranging between 0.94 and 9.07 and the 95% CI of 0.92-89.9 when compared with health facility DOT, self-administered treatment and community health care worker DOT. Of these four, one study in Tanzania reported a low risk ratio to treatment success with the other three having a risk ratio of above one.(50)(48)(40)(41)

### Short Message Service Reminders (SMS)

Four RCTs(15) (16) (68)(18) evaluating SMS as medication reminders showed no statistically significant effect on treatment completion, when compared with the local standard of TB care. In three of these from the low and middle income countries of Cameron, Argentina and Pakistan conducted between the years of 2013 and 2018 (15) (68)(18), the risk ratios for completion, success or cure ranged from 1.0–1.45, and the 95% CI fell within these values in all three trials. One multinational RCT conducted in Spain, Hong Kong, US and South Africa reported 76.4% vs 85.4% (95% CI:71.3-80.8%) treatment completion when comparing SMS with facility DOT.(16)This results shows that SMS DOT is an effective intervention to successful treatment outcomes.

### Models economic viability/cost effectiveness

Cost effectiveness was determined by dividing the total cost for each model by the treatment success rate at 6 months. Final costs were estimated and costs per patient cured were compared with the aim to determine the average cost and the marginal or incremental cost for an additional unit of health benefit when choosing between two models, the incremental cost effectiveness ratio and average cost effectiveness was used. The incremental cost effectiveness ratio was defined as difference in total costs between intervention and control divided by the difference in the number of patients with sputum conversion and who completed treatment between intervention and control.

### Video observed treatment (VOT)

Four observational studies in high income countries of Australia and United States of America between 2012 and 2021 evaluated the cost effectiveness of this model, Three of these have shown an incremental cost-savings ranging between $1391 and $2,226 when comparing VOT with using in-person DOT and VOT was therefore the preferred cost-effective option(35)(36)(43). Another retrospective cohort design in Australia comparing patients receiving direct observation by home videophone with patients receiving this service in person have shown the incremental cost-effectiveness ratio (ICER) to be AUD$1.32 (95% CI: $0.51 - $2.26) per extra day of successful observation with VOT.(44) The video service used less staff time, and became dominant if implemented on a larger scale and/or with decreased technology costs

### Electronic medication monitor (EMM)

Two observational studies in low-and middle-income countries of South Africa and Morrocco in 2012 and 2021 respectively evaluated cost effectiveness under this model. One study in South Africa comparing the costs and health outcomes of the DOTS-SIMpill cohort with DOTS-only controls has shown a positive return on investment (ROI of 23% over the 5-year period) for the DOTS-SIMpill cohort based on improved health outcomes and reduced average cost per patient.(30) Another study conducted in Morocco evaluating the costs and cost-effectiveness of a Medication Event Monitoring System (using a smart pillbox with a web-based medication monitoring system)for Tuberculosis Management against standard of care (SoC) have shown the ICER of $434/DALY averted for managing drug-susceptible TB patients by MEMS relative to SoC thus MEMS is considered cost-effective in Morocco.(47)

### Community health care worker Directly observed therapy (CHWDOT)

Three randomized control trials evaluating the cost effectiveness of community health care worker DOT in low and middle income countries of Ethiopia, Pakistan and Nigeria between 2002 and 2015 have reported in favour of community health care worker DOT with incremental cost effectiveness ratio ranging between 16.3 and 410 US$ compared to health facility based and self-administered DOT.(19)(14)(25) The community health worker became the most cost effective approach.

### Family-member Directly observed therapy (FDOT)

Two observational studies in low-and middle-income countries of Brazil and Thailand conducted in 2011 and 2012 evaluated family DOT model cost effectiveness, they all reported in favour of the model when compared with community health worker DOT and health care worker and Self-administered DOT. A study in Brazil have shown the guardian supervised/family DOT cost on average to be US$398 per patient cured. This figure was US$ 260(39%) lower than its equivalent for CHW supervised DOT (US$657) resulting in saving of US$1.0095 per additional patient cured.(50) A study in Thailand have reported a cost savings associated with family-member DOT (−I$9 million [95% uncertainty interval −I$12 million to −I$5 million]) with 9400 DALYs averted, ICER I$1100 dominant to I$1300 indicating that family-member DOT is a cost-saving intervention.(37)

### Short Message Service Reminders

Two observational studies conducted in low-and middle-income countries of Thailand and Brazil in 2012 and 2018 have shown SMS intervention to be cost effective when compared with and SAT. An economic study in Thailand on SMS use in TB care in demonstrated an incremental cost-effectiveness ratio of 350 ‘international dollars’ per disability adjusted life year (37). Another decision analysis model developed to simulate cohorts who initiate TB treatment in Brazil compared SMS intervention have shown that, among persons from the general population with latent TB infection, SMS was the most cost-effective intervention against SAT with the incremental cost (95% UR) of USD 164 (USD 29 saving to USD 362 cost) per DALY averted and USD 814 (USD 137 saving to USD 1781 cost) per TB case prevented. SMS cost USD 1000 per DALY averted and USD 4483 per TB case prevented compared to SAT(52).

## Discussion

Community TB care interventions are increasingly used to support TB treatment in diverse settings globally. This analysis is set to examine the potential cost and impact of various Community TB care interventions/models as applied under programme conditions globally. It provides preliminary insights into the potential impact and cost of several approaches to TB treatment support in this context. We estimated effectiveness and substantial cost savings with VOT, medication monitors, community health worker DOT, family DOT and SMS including savings to patients and their families, compared to other treatment options. While evidence remains incomplete, and generalisability limited, the studies reviewed suggest these interventions may improve efficiency, save money and reduce burden on patients and healthcare workers.

A cohort study in Moldova reported a protective facility DOT over VOT whereas studies in the USA, UK and Australia suggested that treatment outcomes were improved compared with in-person DOT, with markedly reduced health system costs. These results suggest that VOT is ineffective in improving treatment success in the low and income countries but more effective in the high-income countries. This could be mainly due to accessibility of digital platforms in developing countries posing a challenge for model efficacy. Therefore, until VOT becomes cheaper, it will probably be substantially less cost-effective for supporting treatment in low income settings while in settings where digital solutions cost less and/or are easier to implement and use than the standard of care, VOT may be a beneficial alternative. (52)(70)

Electronic medication Monitoring System improved the TB treatment success rates in this review. Over time, the EMM group showed a higher treatment success rate and cost saving compared to the standard conventional TB treatment over a six-month period. EMM is expected to contribute to the effectiveness (treatment outcomes and cost) of the TB case management strategy based on its convenient and effective monitoring of medication in low-and middle-income countries

This review finding has shown that community-based DOT produced outcomes that were equivalent and or superior to the other treatment options for TB patients suggesting that CHW can effectively dispense anti-TB medication and community participation should be encouraged. The implication of this in high TB burden settings is that community-based TB treatment is an effective and viable option that can supplement other modes of treatment delivery. Furthermore, community-based TB treatment delivery has been found to be cost-effective, and it is a low-cost technology that can easily be adapted to diverse areas of need and appropriate CHW recruited according to availability in each contextual setting.

Family-supervised DOTS was more effective and less costly than other forms of DOT delivery. Implementation of Family -supervised DOTS exceeded the quality of patient outcomes from other treatment options. In cost-effectiveness parlance, the results indicate that guardian-supervised DOTS was the dominant strategy. Possible explanation is that direct observation by family members incurs little inconvenience and negligible costs to the patient, as patients, once well enough, are not constrained in continuing their normal work. In addition, there is less potential for stigma. (23)

This systematic review shown paucity of high-quality evidence concerning effect of mobile-phone messaging on anti-TB treatment success, All the studies in this review were all from low-and middle-income countries. Mobile-phone messaging showed a modest effect in improving TB treatment success and cost saving. Results from this review concurs with evidence from meta-analyses of RCTs in other disease settings which have shown a positive effect of two-way SMS on treatment outcomes, which suggests that compared with DOT, SMS reminding regularly could significantly increase the patients’ successful TB treatment.(69) The most likely reason is that besides daily drug intake schedules, the SMS group patients could receive extra frequent prompts and health information, which gradually propel patients to practicing good habits and health awareness.(68)

## Limitations

Our review has several limitations. We focused on quantitative comparisons of clinical outcomes, as these are fundamental to the evidence base. For this reason, we have not provided a detailed review of studies which focused exclusively on qualitative assessments. Given the marked heterogeneity of study designs, endpoints, and settings, we were unable to pool the estimates of effect, and could only summarise findings as reported from each of the studies. The other weakness of this study was the necessity of using a retrospective cohort comparison between the intervention and control groups. Whilst matching was undertaken for the available data, other confounding factors may have existed and the effect of these is unknown.

## Conclusions

In conclusion, this review illustrates that community-based TB interventions such as VOT, CHWDOT, FDOT, SMS and EMMs plays a successful role in improving the treatment success rate in low-and middle-income countries. Scarce resources can be conserved by managing larger numbers of TB patients with the same number of staff thus cost effective. Moreover, community-based DOT can substantially bolster efficiency and convenience for patients and providers thus saving costs and improving clinical outcomes.

## Data Availability

All relevant data are within the manuscript and its Supporting Information files.

## Supporting information

S1 Fig 1: Preferred Reporting Items for Systematic and Meta Analyses flow diagram. (Adapted from Moher et al).(11)

S2 Fig 2: Summary of articles showing model of implementation and country classification by economic level.

S3 Fig 3. The risk of bias assessment of included papers using ROB 2 tool for randomized studies.

S4 Fig 4. The risk of bias assessment of included papers using ROB 2 tool for cluster-randomized studies.

S5 Fig 5. The risk of bias assessment of included papers using ROBINS-I tool for non-randomized studies.

S1 Table 1: Data extraction form

S1 Table 2: General information for included studies on Community Based TB interventions and their impact on treatment outcomes (28 studies).

S3 Table 3: General information for included studies on Community Based TB interventions and their cost effectiveness (12 studies)

S4 Table 4: List of excluded studies along with reasons for exclusion Prisma checklist

### Funding

This research was undertaken as part of a PhD funded by the University of KwaZulu Natal, Durban, South Africa (www.ukzn.ac.za). The funders had no role in study design, data collection and analysis, decision to publish, or preparation of the manuscript

### Competing Interests

The authors declare that no competing interests exist.

## Notes

### Competing Interest Statement

The authors have declared no competing interest.

### Author Declarations

Ethical review was not needed since it was systematic review

## References

1. World Health Organization. Global tuberculosis report 2019. World Health Organization. License: CC BY-NC-SA 3.0 IGO [Internet]. 2019. Available from: https://apps.who.int/iris/handle/10665/329368

2. World Health Organisation. Global tuberculosis report 2021 World Health Organization. Licence: CC BY-NC-SA 3.0 IGO, Geneva [Internet]. 2021. Available from: https://www.who.int/publications/i/item/9789240037021

3. World Health Organization. Implementing the end TB strategy: the essentials. World Health Organization [Internet]. 2015. Available from: https://apps.who.int/iris/handle/10665/206499

4. World Health Organization. Global tuberculosis report 2020. World Health Organization.License: CC BY-NC-SA 3.0 IGO [Internet]. 2020. Available from: https://apps.who.int/iris/handle/10665/336069

5. Loveday M, Wallengren K, Brust J, Roberts J, Voce A, Margot B, et al. Community-based care vs. centralised hospitalisation for MDRTB patients, KwaZulu-Natal, South Africa. Int J Tuberc Lung Dis. 2015;19(2):163–71.

6. Arshad A, Salam RA, Lassi ZS, Das JK, Naqvi I, Bhutta ZA. Community based interventions for the prevention and control of tuberculosis. Infect Dis Poverty. 2014;3(1).

7. Collin SM, Wurie F, Muzyamba MC, de Vries G, Lönnroth K, Migliori GB, et al. Effectiveness of interventions for reducing TB incidence in countries with low TB incidence: A systematic review of reviews. Eur Respir Rev [Internet]. 2019;28(152). Available from: http://dx.doi.org/10.1183/16000617.0107-2018

8. Higgins JPT, Thomas J, Chandler J, Cumpston M, Li T, Page MJ, et al. Cochrane handbook for systematic reviews of interventions. Cochrane Handbook for Systematic Reviews of Interventions. 2019. 1–694 p.

9. Tricco AC, Lillie E, Zarin W, O’Brien KK, Colquhoun H, Levac D, et al. PRISMA extension for scoping reviews (PRISMA-ScR): Checklist and explanation. Ann Intern Med. 2018;169(7):467–73.

10. Danielle Levac HC, Kelly K O’Brien. Scoping studies: advancing the methodology. Implement Sci. 2010;5(1):1–9.

11. Moher D, Liberati A, Tetzlaff J, Altman DG. Preferred reporting items for systematic reviews and meta-analyses: The PRISMA statement. BMJ. 2009;339(7716):332–6.

12. Sterne JAC, Savović J, Page MJ, Elbers RG, Blencowe NS, Boutron I, et al. RoB 2: A revised tool for assessing risk of bias in randomised trials. BMJ. 2019;366:1–8.

13. Sterne JA, Hernán MA, Reeves BC, Savović J, Berkman ND, Viswanathan M, et al. ROBINS-I: A tool for assessing risk of bias in non-randomised studies of interventions. BMJ. 2016;355:4–10.

14. Adewole OO, Oladele T, Osunkoya AH, Erhabor GE, Adewole TO, Adeola O, et al. A randomized controlled study comparing community based with health facility based direct observation of treatment models on patients’ satisfaction and TB treatment outcome in Nigeria. Trans R Soc Trop Med Hyg. 2015;109(12):783–92.

15. Bediang G, Stoll B, Elia N, Abena JL, Geissbuhler A. SMS reminders to improve adherence and cure of tuberculosis patients in Cameroon (TB-SMS Cameroon): A randomised controlled trial. BMC Public Health. 2018;18(1):1–14.

16. Belknap R, Holland D, Feng PJ, Millet JP, Cayla JA, Martinson NA, et al. Self-administered versus directly observed once-weekly isoniazid and rifapentine treatment of latent tuberculosis infection. Ann Intern Med. 2017;167(10):689–97.

17. Iribarren S, Beck S, Pearce PF, Chirico C, Etchevarria M, Cardinale D, et al. TextTB: A Mixed Method Pilot Study Evaluating Acceptance, Feasibility, and Exploring Initial Efficacy of a Text Messaging Intervention to Support TB Treatment Adherence. Tuberc Res Treat. 2013;2013:1–12.

18. Mohammed S, Glennerster R, Khan AJ. Impact of a daily SMS medication reminder system on tuberculosis treatment outcomes: A randomized controlled trial. PLoS One. 2016;11(11):1–13.

19. Khan MA, Walley JD, Witter SN, Imran A, Safdar N. Costs and cost-effectiveness of different DOT strategies for the treatment of tuberculosis in Pakistan. Health Policy Plan. 2002;17(2):178–86.

20. Story A, Aldridge RW, Smith CM, Garber E, Hall J, Ferenando G, et al. Smartphone-enabled video-observed versus directly observed treatment for tuberculosis: a multicentre, analyst-blinded, randomised, controlled superiority trial. Lancet [Internet]. 2019;393(10177):1216–24. Available from: http://dx.doi.org/10.1016/S0140-6736(18)32993-3

21. Walley JD, Khan MA, Newell JN, Khan MH. Effectiveness of the direct observation component of DOTS for tuberculosis: A randomised controlled trial in Pakistan. Lancet. 2001;357(9257):664–9.

22. Wandwalo E, Kapalata N, Egwaga S, Morkve O. Effectiveness of community-based directly observed treatment for tuberculosis in an urban setting in Tanzania: A randomised controlled trial. Int J Tuberc Lung Dis. 2004;8(10):1248–54.

23. Wright J, Walley J, Philip A, Pushpananthan S, Dlamini E, Newell J, et al. Direct observation of treatment for tuberculosis: A randomized controlled trial of community health workers versus family members. Trop Med Int Heal. 2004;9(5):559–65.

24. Zwarenstein M, Schoeman JH, Vundule C, Lombard CJ, Tatley M. A randomised controlled trial of lay health workers as direct observers for treatment of tuberculosis. Int J Tuberc Lung Dis. 2000;4(6):550–4.

25. Datiko DG, Lindtjørn B. Cost and cost-effectiveness of treating smear-positive tuberculosis by health extension workers in Ethiopia: An ancillary cost-effectiveness analysis of community randomized trial. PLoS One. 2010;5(2):1–7.

26. Sylla Thiam, Andrea M LeFevre, Fatoumata Hane, Alimatou Ndiaye, Fatoumata Ba, Katherine L Fielding, Moustapha Ndir CL. Effectiveness of a Strategy to Improve Adherence to Tuberculosis Treatment in a Resource-Poor Setting A Cluster Randomized Controlled Trial. JAMA [Internet]. 2007;297(4):2003–5. Available from: oi: 10.1001/jama.297.4.380.

27. Lwilla F, Schellenberg D, Masanja H, Acosta C, Galindo C, Aponte J, et al. Evaluation of efficacy of community-based vs. institutional-based direct observed short-course treatment for the control of tuberculosis in Kilombero district, Tanzania. Trop Med Int Heal. 2003;8(3):204–10.

28. Newell JN, Baral SC, Pande SB, Bam DS, Malla P. Family-member DOTS and community DOTS for tuberculosis control in Nepal: Cluster-randomised controlled trial. Lancet. 2006;367(9514):903–9.

29. Adatu F, Odeke R, Mugenyi M, Gargioni G, McCray E, Schneider E, et al. Implementation of the DOTS strategy for tuberculosis control in rural Kiboga District, Uganda, offering patients the option of treatment supervision in the community, 1998-1999. Int J Tuberc Lung Dis. 2003;7(9 SUPPL. 1):1998–9.

30. Broomhead S, Mars M. Retrospective return on investment analysis of an electronic treatment adherence device piloted in the Northern Cape Province. Telemed e-Health. 2012;18(1):24–31.

31. Cavalcante SC, Soares ECC, Pacheco AGF, Chaisson RE, Durovni B, Oliveira J, et al. Community DOT for tuberculosis in a Brazilian favela: Comparison with a clinic model. Int J Tuberc Lung Dis. 2007;11(5):544–9.

32. Chuck C, Robinson E, Macaraig M, Alexander M, Burzynski J. Enhancing management of tuberculosis treatment with video directly observed therapy in New York City. Int J Tuberc Lung Dis. 2016;20(5):588–93.

33. Dobler CC, Korver S, Batbayar O, Oyuntsetseg S, Tsolmon B, Wright C, et al. Success of community-based directly observed anti-tuberculosis treatment in Mongolia. Int J Tuberc Lung Dis. 2015;19(6):657–62.

34. Doltu S, Ciobanu A, Sereda Y, Persian R, Ravenscroft L, Kasyan L, et al. Short and long-term outcomes of video observed treatment in tuberculosis patients, the Republic of Moldova. J Infect Dev Ctries. 2021;15(91):17S–24S.

35. Fekadu G, Jiang X, Yao J, You JHS. Cost-effectiveness of video-observed therapy for ambulatory management of active tuberculosis during the COVID-19 pandemic in a high-income country. Int J Infect Dis [Internet]. 2021;113:271–8. Available from: https://doi.org/10.1016/j.ijid.2021.10.029

36. Holzman SB, Zenilman A, Shah M. Advancing patient-centered care in tuberculosis management: A mixed-methods appraisal of video directly observed therapy. Open Forum Infect Dis. 2018;5(4).

37. Hunchangsith P, Barendregt JJ, Vos T, Bertram M. Cost-effectiveness of various tuberculosis control strategies in Thailand. Value Heal [Internet]. 2012;15(1 SUPPL.):S50–5. Available from: http://dx.doi.org/10.1016/j.jval.2011.11.006

38. Kironde S, Kahirimbanyi M. Community participation in primary health care (PHC) programmes: lessons from tuberculosis treatment delivery in South Africa. Afr Health Sci. 2002;2(1):16–23.

39. David K. Mafigiri,Janet W. Mcgrath CCW. Task shifting for tuberculosis control: A qualitative study of community-based directly observed therapy in urban Uganda. Glob Public Health. 2013;7(3):270–84.

40. Mhimbira F, Hella J, Maroa T, Kisandu S, Chiryamkubi M, Said K, et al. Home-based and facility-based directly observed therapy of tuberculosis treatment under programmatic conditions in urban Tanzania. PLoS One. 2016;11(8):1–13.

41. Mathema B, Pande SB, Jochem K, Houston RA, Smith I, Bam DS, et al. Tuberculosis treatment in Nepal: A rapid assessment of government centers using different types of patient supervision. Int J Tuberc Lung Dis. 2001;5(10):912–9.

42. Park S, Sentissi I, Gil SJ, Park WS, Oh BK, Son AR, et al. Medication event monitoring system for infectious tuberculosis treatment in Morocco: A retrospective cohort study. Int J Environ Res Public Health. 2019;16(3).

43. Salcedo J, Rosales M, Kim JS, Nuno D, Suen S, Chang AH. Cost-effectiveness of artificial intelligence monitoring for active tuberculosis treatment: A modeling study. PLoS One [Internet]. 2021;16(July):1–15. Available from: http://dx.doi.org/10.1371/journal.pone.0254950

44. Wade VA, Karnon J, Eliott JA, Hiller JE. Home Videophones Improve Direct Observation in Tuberculosis Treatment: A Mixed Methods Evaluation. PLoS One. 2012;7(11):1–13.

45. Wang N, Shewade HD, Thekkur P, Zhang H, Yuan Y, Wang X, et al. Do electronic medication monitors improve tuberculosis treatment outcomes? Programmatic experience from China. PLoS One [Internet]. 2020;15(11 November):1–12. Available from: http://dx.doi.org/10.1371/journal.pone.0242112

46. Wang N, Guo L, Shewade HD, Thekkur P, Zhang H, Yuan YL, et al. Effect of using electronic medication monitors on tuberculosis treatment outcomes in China: a longitudinal ecological study. Infect Dis Poverty [Internet]. 2021;10(1):1–9. Available from: https://doi.org/10.1186/s40249-021-00818-3

47. Yang J, Kim H, Park S, Green N, Kim Y. Cost-Effectiveness of a Medication Event Monitoring System for Tuberculosis Management in Morocco. Res Sq [Internet]. 2021;1– 11. Available from: doi: 10.21203/rs.3.rs-970723/v1.

48. Akkslip S, Rasmithat S, Maher D, Sawert H. Direct observation of tuberculosis treatment by supervised family members in Yasothorn Province, Thailand. Int J Tuberc Lung Dis. 1999;3(12):1061–5.

49. Prado TN do, Rajan J V., Miranda AE, Dias E dos S, Cosme LB, Possuelo LG, et al. Clinical and epidemiological characteristics associated with unfavorable tuberculosis treatment outcomes in TB-HIV co-infected patients in Brazil: a hierarchical polytomous analysis. Brazilian J Infect Dis. 2017;21(2):162–70.

50. Thiago Nascimento do Prado, Nikolas Wada, Leticia Molino Guidoni, Jonathan E. Golub, Reynaldo Dietze ELN. Cost-effectiveness of community health worker versus home-based guardians for directly observed treatment of tuberculosis in Vitória, Espírito Santo State, Brazil. Cad Saúde Pública [Internet]. 2011;27(5):944–52. Available from: https://doi.org/10.1590/S0102-311X2011000500012

51. Sesay ML. Patient Characteristics and Treatment Outcomes Among Tuberculosis Patients in Sierra Leone. walden university; 2017.

52. Nsengiyumva NP, Mappin-Kasirer B, Oxlade O, Bastos M, Trajman A, Falzon D, et al. Evaluating the potential costs and impact of digital health technologies for tuberculosis treatment support. Eur Respir J [Internet]. 2018;52(5). Available from: http://dx.doi.org/10.1183/13993003.01363-2018

53. Dudley L, Azevedo V, Grant R, Schoeman JH, Dikweni L, Mahers D. Evaluation of community contribution to tuberculosis control in Cape Town, South Africa. Int J Tuberc Lung Dis. 2003;7(9 SUPPL. 1):48–55.

54. Becx-Bleumink M, Wibowo H, Apriani W, Vrakking H. High tuberculosis notification and treatment success rates through community participation in central Sulawesi, Republic of Indonesia. Int J Tuberc Lung Dis. 2001;5(10):920–5.

55. Singh AA, Parasher D, Shekhavat GS, Sahu S, Wares DF, Granich R. Effectiveness of urban community volunteers in directly observed treatment of tuberculosis patients: A field report from Haryana, North India. Int J Tuberc Lung Dis. 2004;8(6):800–2.

56. Barker RD, Millard FJC, Nthangeni ME. Unpaid community volunteers - Effective providers of directly observed therapy (DOT) in rural South Africa. South African Med J. 2002;92(4):291–4.

57. Garfein RS, Liu L, Cuevas-Mota J, Collins K, Muñoz F, Catanzaro DG, et al. Tuberculosis treatment monitoring by video directly observed therapy in 5 health districts, California, USA. Emerg Infect Dis. 2018;24(10):1806–15.

58. DeMaio J, Schwartz L, Cooley P, Tice A. The application of telemedicine technology to a directly observed therapy program for tuberculosis: A pilot project. Clin Infect Dis. 2001;33(12):2082–4.

59. Holzschuh EL, Province S, Johnson K, Walls C, Shemwell C, Martin G, et al. Use of Video Directly Observed Therapy for Treatment of Latent Tuberculosis Infection — Johnson County, Kansas, 2015. MMWR Morb Mortal Wkly Rep. 2017;66(14):387–9.

60. Denkinger CM, Grenier J, Stratis AK, Akkihal A, Pant-Pai N, Pai M. Mobile health to improve tuberculosis care and control: A call worth making. Int J Tuberc Lung Dis. 2013;17(6):719–27.

61. Kabongo D, Mash B. Effectiveness of home-based directly observed treatment for tuberculosis in Kweneng West subdistrict, Botswana. African J Prim Heal Care Fam Med. 2010;2(10).

62. Zvavamwe Z, Ehlers VJ. Experiences of a community-based tuberculosis treatment programme in Namibia: A comparative cohort study. Int J Nurs Stud. 2009;46(3):302–9.

63. Niazi AD, Al-Delaimi AM. Impact of community participation on treatment outcomes and compliance of DOTS patients in Iraq. East Mediterr Heal J. 2003;9(4):709–17.

64. Garfein RS, Collins K, Muñoz F, Moser K, Cerecer-Callu P, Raab F, et al. Feasibility of tuberculosis treatment monitoring by video directly observed therapy: A binational pilot study. Int J Tuberc Lung Dis. 2015;19(9):1057–64.

65. Ollé-Goig JE, Alvarez J. Treatment of tuberculosis in a rural area of Haiti: Directly observed and non-observed regimens. The experience of Hôpital Albert Schweitzer. Int J Tuberc Lung Dis. 2001;5(2):137–41.

66. Dave PV, Shah AN, Nimavat PB, Modi BB, Pujara KR, Patel P, et al. Direct observation of treatment provided by a family member as compared to non-family member among children with new tuberculosis: A pragmatic, non-inferiority, cluster-randomized trial in Gujarat, India. PLoS One. 2016;11(2):1–14.

67. Manders AJE, Banerjee A, Van den Borne HW, Harries AD, Kok GJ, Salaniponi FML. Can guardians supervise TB treatment as well as health workers? A study on adherence during the intensive phase. Int J Tuberc Lung Dis. 2001;5(9):838–42.

68. Fang XH, Guan SY, Tang L, Tao FB, Zou Z, Wang JX, et al. Effect of short message service on management of pulmonary tuberculosis patients in Anhui Province, China: A prospective, randomized, controlled study. Med Sci Monit. 2017;23:2465–9.

69. Gashu KD, Gelaye KA, Mekonnen ZA, Lester R, Tilahun B. Does phone messaging improves tuberculosis treatment success? A systematic review and meta-analysis. BMC Infect Dis. 2020;20(1):1–13.

70. Ngwatu BK, Nsengiyumva NP, Oxlade O, Mappin-Kasirer B, Nguyen NL, Jaramillo E, et al. The impact of digital health technologies on tuberculosis treatment: A systematic review. Eur Respir J [Internet]. 2018;51(1). Available from: http://dx.doi.org/10.1183/13993003.01596-2017

